# Deep Learning-Derived Myocardial Strain

**DOI:** 10.1101/2022.03.16.22272374

**Authors:** Alan C. Kwan, Márton Tokodi, Ishan Jain, Ernest Chang, John Theurer, Xiu Tang, Nadia Francisco, Francois Haddad, David Liang, Neal Yuan, Béla Merkely, Robert Siegel, Susan Cheng, Attila Kovács, David Ouyang

**Author notes:** **Correspondence:** Mail: 127 S. San Vicente Blvd. AHSP Pavilion Suite A3100, Los Angeles, CA 90048.

## Abstract

**Background:** Echocardiographic strain measurements require extensive operator experience and have significant inter-vendor variability. This study sought to develop an automated deep learning strain (DLS) analysis pipeline and validate its performance both externally and prospectively.

**Methods:** The DLS pipeline takes blood pool semantic segmentation results from the EchoNet-Dynamic network and derives longitudinal strain from the frame-by-frame change in the length of the left ventricular endocardial contour. The pipeline was developed using 7,465 echocardiographic videos, with preprocessing steps optimized to determine the change in endocardial length from systole to diastole. It was evaluated on a large external retrospective dataset and was prospectively compared with manual within-patient acquisition of repeated measures by two experienced sonographers and two separate vendor speckle-tracking methods on different machines.

**Results:** In the external validation set, the DLS method maintained moderate agreement (intraclass correlation coefficient (ICC) 0.58, p<0.001) with a bias of -2.33% (limits of agreement -10.61 to 5.93%). The absolute difference in measurements was independent of subjective image quality (ß: 0.12, SE: 0.10, p=0.21). Compared to readers on repeated measures, our method has reduced variability (standard deviation: 1.35 vs. 2.55%) and better inter-vendor agreement (ICC: 0.45 vs. 0.29).

**Conclusions:** The DLS measurement provides lower variance than human measurements and similar quantitative results. The method is rapid, consistent, vendor-agnostic, publicly released, and robust across a wide range of imaging qualities.

## INTRODUCTION

Myocardial strain can be used to identify subtle signatures of cardiac dysfunction.^1^ It has specific applications in the longitudinal monitoring of cardiotoxic therapies as well as in diagnosing cardiomyopathies.^2^ Even compared to conventional measures of left ventricular (LV) function such as ejection fraction, longitudinal strain has incremental benefits for predicting cardiovascular outcomes.^3-5^ Strain is formally defined as the change in the length (L) of a myocardial segment relative to its initial diastolic length (L_0_): strain = (L-L_0_)/ L_0_.^6^ However, the method of measuring the length of a myocardial segment can significantly vary across manufacturers which may impact the downstream assessment of strain and hamper the comparison of strain values across different vendors.^7^ In echocardiography, speckle tracking is the primary method of obtaining longitudinal strain.^1^ Nonetheless, the application of this method requires significant experience^8^, and it exhibits significant inter-/intra-vendor variability, limiting the ability to compare assessments over time and between different locations.^9, 10^ Creation of an open-source, vendor-agnostic method to retrospectively measure longitudinal strain from standard B-mode images would greatly improve post-hoc research applications and benefit patient care by improving access and ability to more directly compare between images captured by different sites, operators, and vendors.

In this manuscript, we present an open-source deep learning-based solution to quantify LV global longitudinal strain (GLS) and evaluate its performance against manual segmentation-based strain measurements in both internal and external validation. We hypothesized that our method of deep learning strain (DLS) assessment could provide consistent and robust strain measurements in an automated fashion, similar to standard speckle tracking echocardiography, but without the need for manual measurements and expertise.

## METHODS

### Study Design

We adapted EchoNet-Dynamic, a deep learning-based segmentation network tailored for analyzing echocardiograms,^11^ to identify the endocardial-blood pool interface across the cardiac cycle using apical 4-chamber (A4C) view echocardiographic videos. GLS was computed using an automated pipeline based on mathematical analysis of the boundaries of the LV across the cardiac cycle. The strain methodology was applied retrospectively over a large external database with 3D echocardiography-derived LV GLS to assess for agreement. The strain methodology was then prospectively compared to standard 2D speckle tracking strain with repeated within-patient measurements from two sonographers using two separate echocardiography vendors to determine inter- and intra-measurement variability. The study protocol was approved by Stanford University, Semmelweis University (#190/2020), and Cedars Sinai Institutional Review Boards.

### Datasets

EchoNet-Dynamic was trained on 7,465 A4C view echocardiographic videos of 7,465 unique patients from Stanford Healthcare and validated internally on an additional 2,565 videos. The dataset is publicly available at https://echonet.github.io/dynamic/. The DLS algorithm was then tested on an external cohort of 813 unique patients with 2,454 A4C videos from Semmelweis University. To compare the intra-provider variation of our DLS method with that of the conventional strain algorithms, 43 unique patients were prospectively scanned four times per patient by two experienced sonographers on two different vendor machines.

### Deep Learning Strain Algorithm

The DLS algorithm builds upon the foundation of a previously validated semantic segmentation model, which identifies the LV area throughout the cardiac cycle.^12^ From the identification of the LV, the algorithm calculates longitudinal strain using input echocardiogram videos by 1) contouring the endocardium around the LV blood pool in the A4C view, 2) identifying and excluding the mitral annular plane, 3) expanding the contour to trace mid-endocardium, 4) measuring the length of the LV in each frame, 5) identifying the longest and shortest lengths in a single cardiac cycle to calculate per-beat strain, and 6) calculating the average the cardiac strain over all of the cardiac cycles within a single video clip (**Figure 1**).

**Figure 1.**
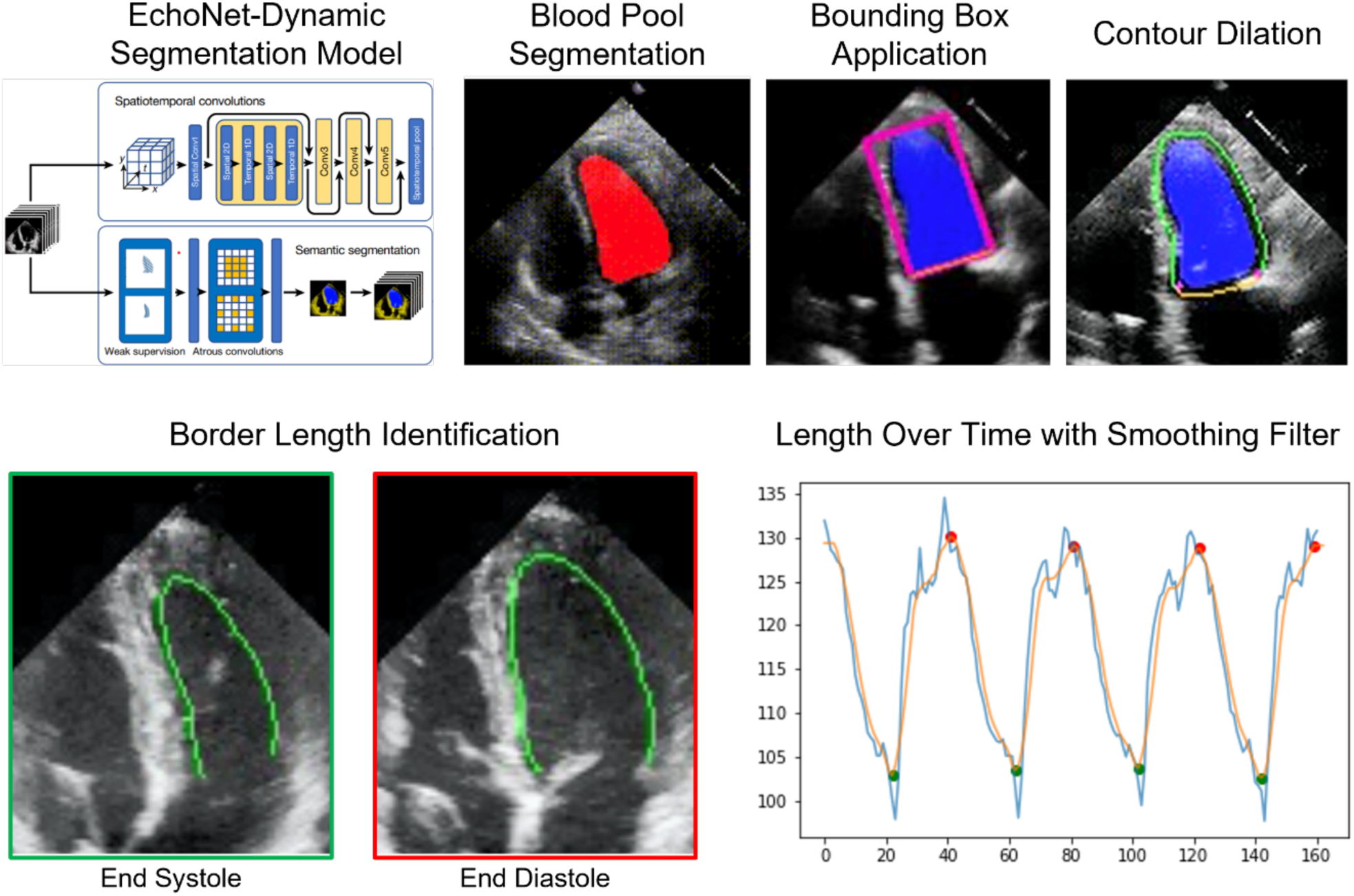
Summary diagram of deep learning-based strain process including passage of blood pool segmentation, bounding box identification for contour identification and dilation. Total border length is measured through the cardiac cycle, with a Savitzky-Golay filter to smooth the contour. The strain is calculated as the mean change in length in the endocardial border within the video clip.

The development of the original semantic segmentation model has been previously described.^11^ Starting with the trained Echonet-Dynamic segmentation model weights, we were able to identify the LV blood pool contour. To exclude the mitral annular plane from the length of the LV endocardium, a bounding box was applied over the blood pool contour to identify the insertion points of the mitral leaflets and exclude the segments of contour crossing the annular plane. Given that the exact boundary of the LV may vary due to the presence of trabeculations, we tested varying degrees of dilation of the border and their effect on reproducibility. The length of this border contour was quantified in each frame. Using Python’s SciPy function scipy.signal.find_peaks,^13^ over a 32-frame sliding window, we identified the maximum and minimum lengths of the LV endocardial tracing, corresponding with systole and diastole. Given the expected smooth contraction and relaxation of the LV, we tested multiple filters length-fame plots to reduce frame-to-frame variance in length across a cardiac cycle. We evaluated multiple filters, including Savitzky-Golay (Savgol), convolve average, moving average, high pass, and low pass. Algorithms were implemented in Python (version 3.9.5, Python Software Foundation, Wilmington, Delaware, USA) using open-source libraries including PyTorch, SciPy, Torchvision, and OpenCV.^14^

### External Validation on a Cohort from a Separate Healthcare System

The DLS pipeline was applied to an external cohort of subjects who underwent clinically-indicated 2D and 3D echocardiographic examination at Semmelweis University between November 2013 and March 2021. The cohort consisted of patients with different cardiac diseases (such as heart failure, cardiomyopathies, valvular heart disease), healthy subjects with no cardiovascular risk factors or established cardiac diseases, and athletes. This external dataset included A4C view echocardiographic videos acquired using various Philips and GE ultrasound systems and 3D echocardiography-derived expert annotations. 3D LV end-diastolic and end-systolic volumes, ejection fraction, LV mass, and GLS were assessed using a commercially available software solution (4D LV-Analysis 3, TomTec Imaging GmbH, Unterschleissheim, Germany). Videos were individually assessed for Turing test failure by identifying clear anatomically erroneous segmentation, and cases with failure were excluded from the analysis.

### Prospective Repeated Measures Analysis

The DLS pipeline was prospectively compared with standard acquisitions in 43 unique patients who received blinded, independent strain analysis by two senior sonographers who independently scanned, analyzed, and interpreted strain across two different machines. Both senior sonographers had advanced cardiac certification and more than 15 years of experience. Each patient was scanned using both Epiq 7C (strain in v2.01) and GE Vivid E95 (strain in QLAB v10.2) ultrasound machines on the same day to acquire a total of 4 acquisitions per patient.

### Statistical Analysis

Continuous variables are expressed as mean (standard deviation), while categorical variables are reported as frequencies and percentages. In the external validation cohort, we compared 3D echocardiography-derived GLS and DLS. In the repeated measures cohort, we evaluated inter-observer, intra-observer, inter-vendor, and cycle-to-cycle variability of the DLS method, and we also compared manual strain versus DLS. Comparisons were made using two-tailed paired T-test, intraclass correlation coefficients (ICC), and Bland-Altman analyses. Statistical analyses were performed in R (version 4.1.1, R Foundation for Statistical Computing, Vienna, Austria).

## RESULTS

The original training cohort included 7,465 patients (70±22 years, 49% women, **Table 1**), with the repeated measures cohort including 43 patients (55±17 years, 38% women).

**Table 1.** Demographics, clinical and echocardiographic characteristics of the training and external validation datasets

|  | Original training dataset | External validation dataset |
| --- | --- | --- |
| Number of patients | 7,465 | 813 |
| Number of videos | 7,465 | 2,454 |
| Age, years (SD) | 70 (22) | 46 (23) |
| Female, n (%) | 3,662 (49) | 298 (37) |
| Heart failure, n (%) | 2,113 (28) | 238 (29) |
| Diabetes mellitus, n (%) | 1,474 (20) | 109 (13) |
| Hypercholesterolemia, n (%) | 2,463 (33) | 224 (28) |
| Hypertension, n (%) | 2,912 (39) | 315 (39) |
| Renal disease, n (%) | 1,475 (20) | 112 (14) |
| Coronary artery disease, n (%) | 1,674 (22) | 103 (13) |
| End diastolic volume, mL (SD) | 91.0 (46.0) | 144.9 (58.7)* |
| End systolic volume, mL (SD) | 43.2 (36.1) | 71.6 (49.0)* |
| Ejection fraction, % (SD) | 55.7 (12.5) | 53.5 (12.3)* |
| Echocardiographic machine |  |  |
| Philips Epiq 7C, n (%) | 4,832 (65) | 340 (14) |
| Philips Epiq 7G, n (%) | 0 (0) | 580 (24) |
| Philips iE33, n (%) | 2,489 (33) | 99 (4) |
| Philips CX50, n (%) | 62 (1) | 0 (0) |
| Philips Epiq 5G, n (%) | 44 (1) | 0 (0) |
| GE Vivid E95, n (%) | 0 (0) | 1,424 (58) |
| Other, n (%) | 38 (1) | 11 (0) |

\*measured with 3D echocardiography
SD – standard deviation

### Pipeline Optimization

Two significant empirical decisions were made while designing the automated pipeline: the degree of dilation and the smoothing filter. For the former, 0-pixel dilation (i.e., no dilation) was compared with 3-pixel and 5-pixel dilation. When dilated with 0 pixels, the mean standard deviation of DLS was 2.57%, whereas 3-pixel and 5-pixel dilation resulted in a mean standard deviation of 2.02% and 2.50%, respectively. For the smoothing filter, Savgol was 1.35%, low-pass was 1.52%, convolve average was 1.40%, and the moving average was 1.89%. Therefore, the Savgol filter with 3-pixel dilation was selected for the final pipeline.

### External Agreement Validation

Out of the 3,812 videos (900 patients, 47±23 years, 38% women) of the external dataset, the DLS method was feasible in 3,488 (91.5%) videos of 881 (97.9%) patients. An additional 1,034 (27.1%) videos were excluded due to anatomically erroneous segmentation. The final external validation dataset comprised 2,454 videos from 917 examinations of 813 patients (46±23 years, 37% women, Table 1). 3D echocardiography-derived GLS ranged from -1.16 to -31.83%, with a mean of -17.79±4.90%, whereas DLS ranged from -6.10 to -38.94%, with a mean of -20.13±5.06%. The ICC between the two measurements was 0.58 (0.37-0.71), p<0.001, with a bias of -2.33% and limits of agreement (LOA) of -10.61 to 5.93 (**Figure 2**). The most significant predictors of the absolute difference between the ground truth and predicted strain values were DLS (ß: -0.37, SE: 0.01, p<0.001), 3D GLS (ß: 0.33, SE: 0.02, p<0.001), 3D LV ejection fraction (ß: -0.09, SE: 0.01, p<0.001), and 3D LV mass (ß: 0.01, SE: 0.001, p<0.001). Subjective image quality (graded on a 1-5 scale) was not significantly associated with the absolute difference in measures (ß: 0.12, SE: 0.10, p=0.21) (**Figure 3**).

**Figure 2.**
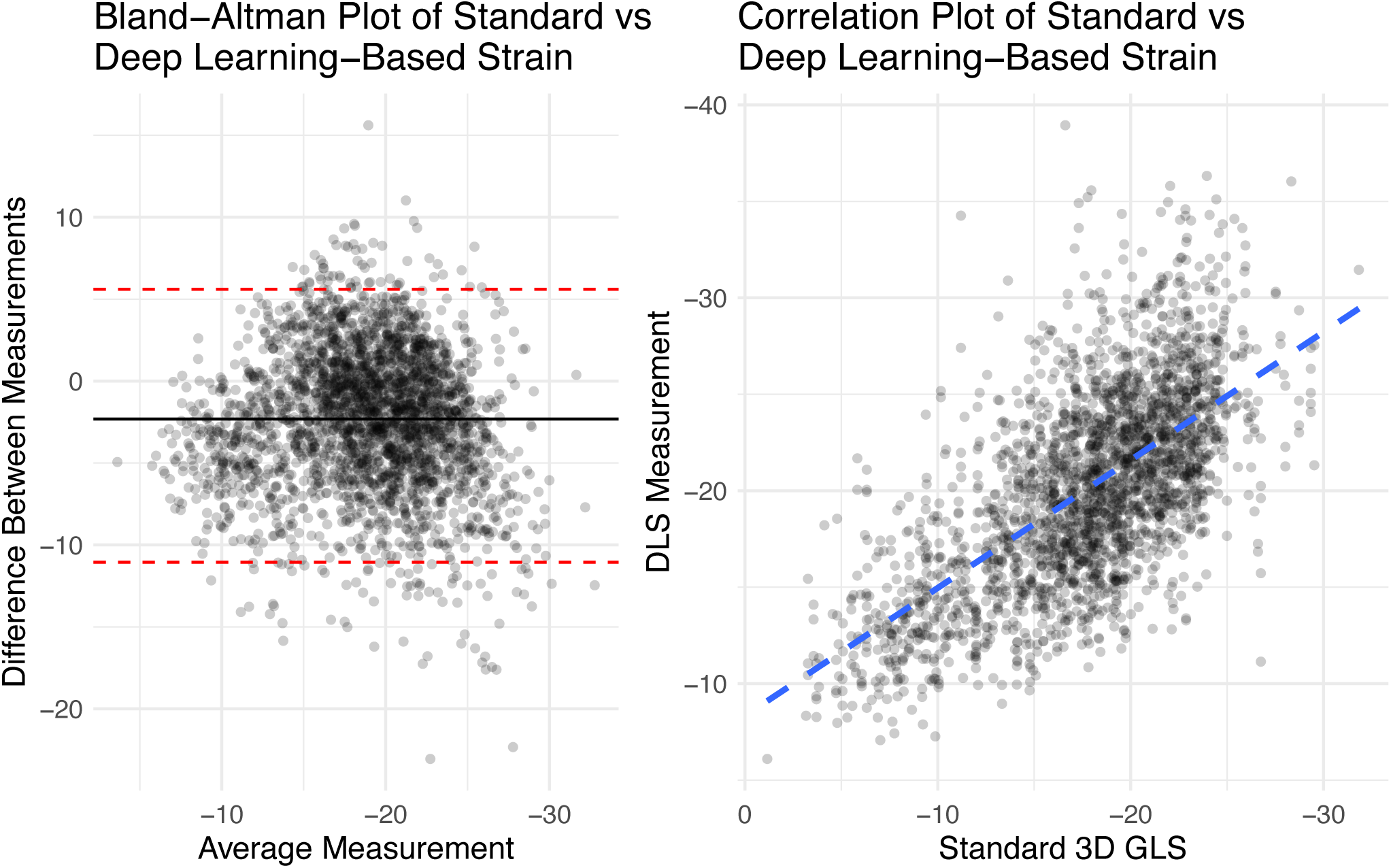
Comparison of 3D echocardiography-based global longitudinal strain and deep learning-derived strain by Bland Altman and Correlation Plots. Deep learning strain was on average less by 2.33%, Limits of Agreement of -10.61 to 5.93%. DLS: Deep learning strain; GLS: Global longitudinal strain.

**Figure 3.**
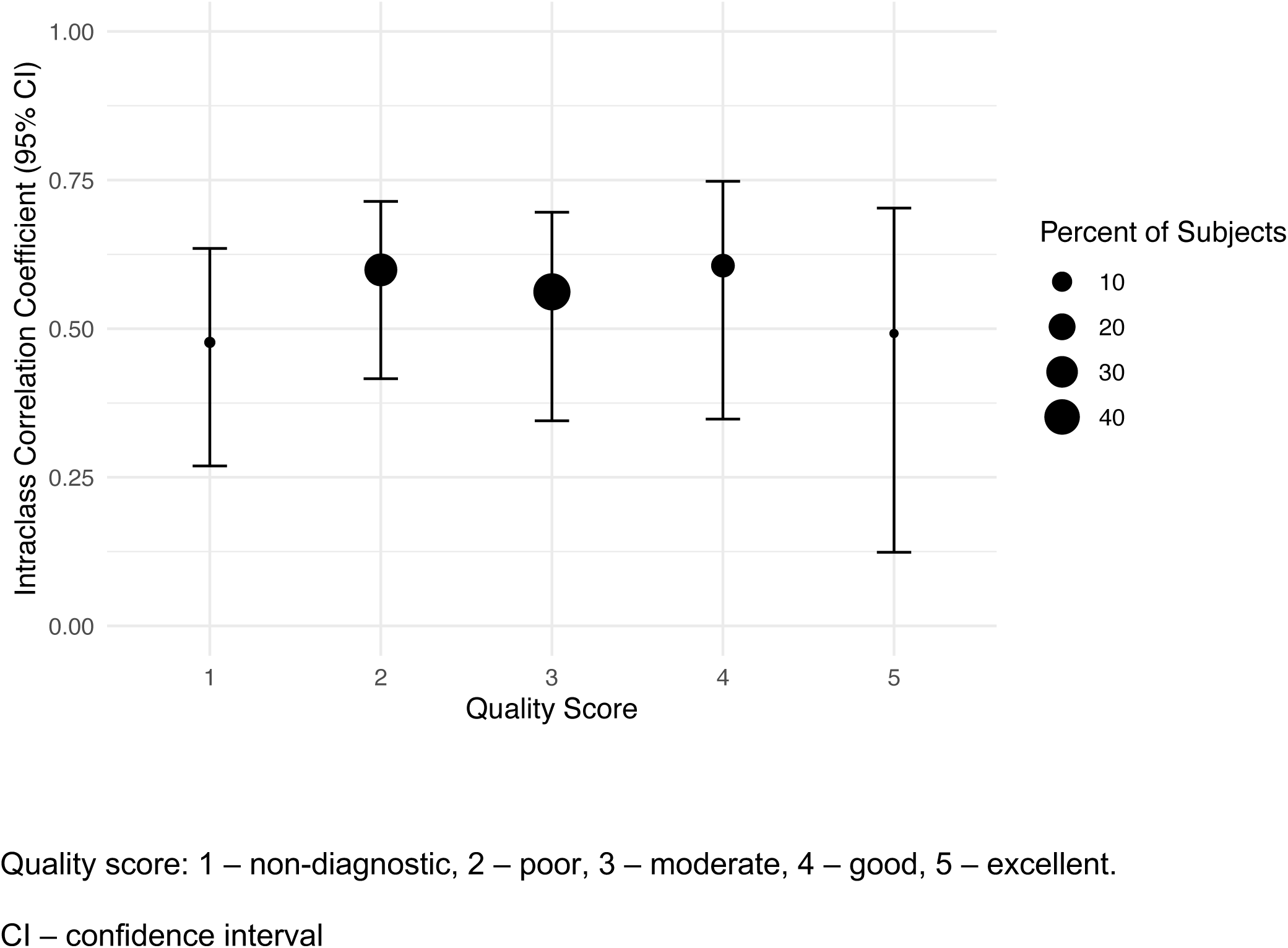
Intraclass correlation coefficient with 95% confidence interval by subjective quality score of the image. A lower quality score indicates worse image quality.

### Prospective Inter-method Variation

In the repeated measures cohort, four manual measurements were performed for each patient: both readers performed strain analysis using the Philips and GE machines. The mean GLS measured by readers 1 and 2 was -18.00±2.64% and -17.90±2.41, respectively. Measurements on Philips and GE had mean and standard deviations of -19.31±1.74% and -16.58±1.39%, respectively. The ICC between readers 1 and 2 was 0.63 (0.48-0.74), p<0.001. The ICC between GE and Philips systems was 0.29 (−0.01-0.53), p=0.03.

In the DLS measurements, the overall cohort had a strain of -15.28±1.35%, with Philips machine measurements having a mean and standard deviation of -14.96±1.42% and GE machine with - 15.61±1.21%, with an ICC of 0.45 (0.18-0.66), p < 0.001. Comparing the combined human versus DLS methods the mean strain was -17.91±2.55% for human and -15.29±1.35% for DLS. The ICC was not significant: 0.28 (−0.09-0.59), p=0.09, with a bias of 2.73% (LOA: -1.63 to 7.08%) (**Figure 4**).

**Figure 4.**
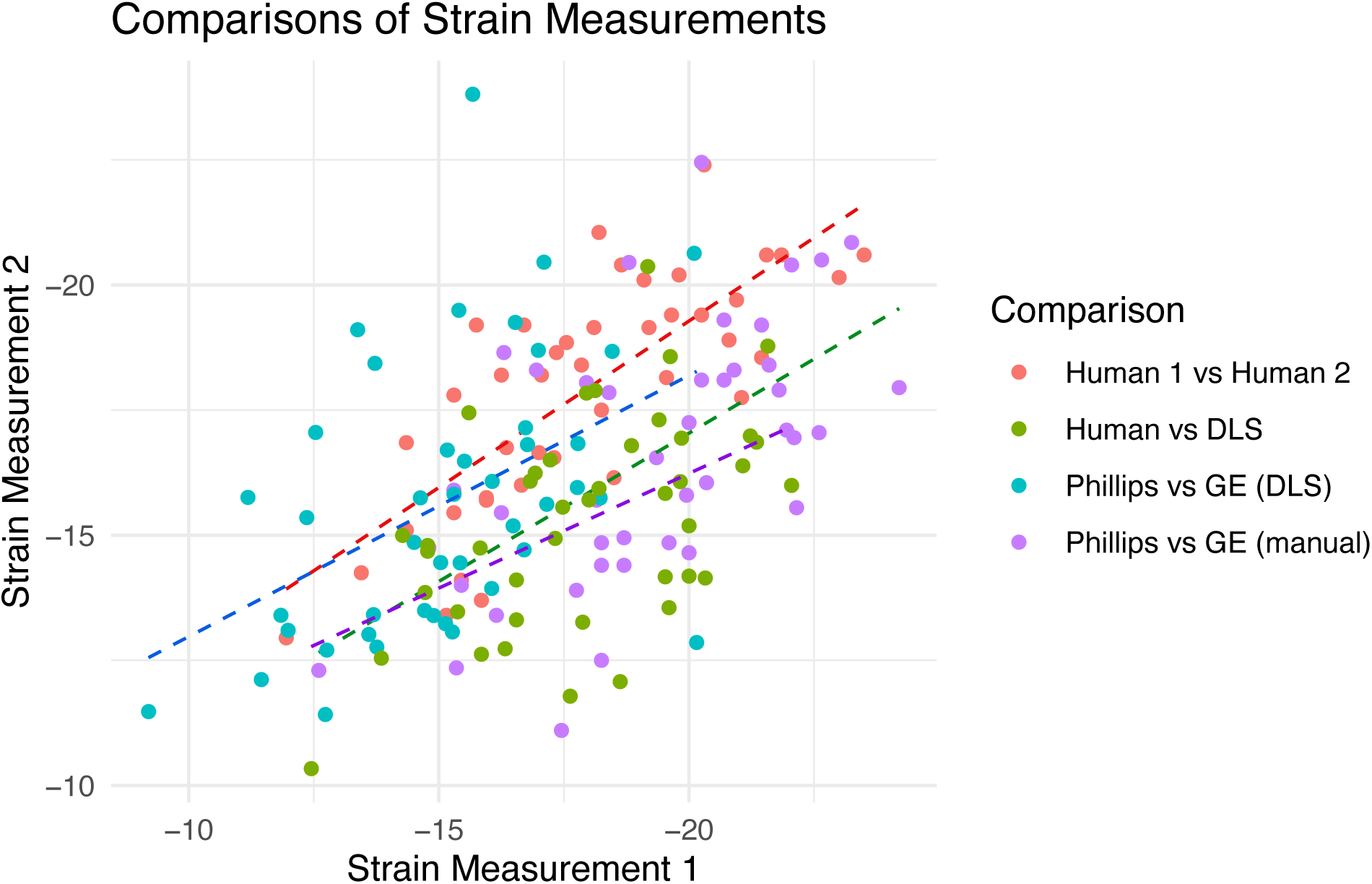
Comparison of different inter-reader methodologies for strain measurement in the prospective cohort. GE: General Electric; DLS: Deep Learning Strain.

Although the standard clinical workflow does not necessitate multi-beat measurement, our deep learning-based method can automatically quantify strain in all cardiac cycles of a given video. In our repeated measures cohort, the mean number of cardiac cycles per video was 3.47. The standard deviation of strain measurements was 1.7%, and the mean difference in the maximum and minimum measurements was 2.6%.

## DISCUSSION

In this manuscript, we presented a vendor-agnostic, open-source, deep learning-based strain analysis tool and compared the predicted strain values to conventional GLS measurement for agreement within and between observers. On an external cohort of patients with 3D echocardiography-derived GLS, our method performed consistently and within the variation seen between different commercial vendors. On a prospective cohort of patients undergoing repeated evaluation by different sonographers and using ultrasound systems of different vendors, our DLS algorithm exhibited lower variability than standard clinical variation by standard deviation, with decreased inter-vendor variability. Notably, between the human and DLS measurements, there was a non-significant agreement, though quantitatively similar to the degree of agreement between human measures on the two separate systems. This may be due to multiple factors, including the smaller size of the prospective cohort, human error, and the inherently different strain measurement methodology. Because our method allows for beat-by-beat assessment of strain, our results demonstrate that there is moderate beat-to-beat variation in strain within a single acquisition, suggesting a lower bound in the precision of strain.

The reference range of different strain methods depends on each commercially available machine and its strain analysis package, which can be limiting given the black-box nature of those software solutions. Our DLS algorithm produces comparable strain values within the reference range suggested by professional societies for normal patients and is open-source and fully automated, allowing for adaptation, iterative improvement, and easy establishment of normal ranges using local data. Given the lack of algorithmic transparency with speckle tracking packages, we show the DLS methodology has enhanced reproducibility, less variance between vendors, and has less dependence on image quality. Additionally, the open-source methods of our technique and development using publicly-available datasets may improve understandability over proprietary methods.

Previous studies have applied deep learning analyses in echocardiography and cardiac magnetic resonance imaging (MRI) for automated strain analysis. Salte et al. passed echocardiographic images through classification networks for view and cardiac phase, then a segmentation network to define the myocardium, then an optical flow network to assess strain.^15^ Given the application of optical flow, similar to speckle tracking, this technique will depend on high resolution and fidelity image quality. Deep learning approaches to semantic segmentation were shown to be robust to even poor video quality,^11^ and could potentially measure strain in low-quality clinical videos where speckle-tracking or optical flow fails. Early application in cardiac MRI for measurement of strain using deep learning in the UK Biobank has been shown.^16^ Given our model’s dependence on semantic segmentation to identify the interface of the blood pool and myocardium, feature-tracking MRI techniques to calculate strain may be more similar to our approach than tagging or even speckle tracking. While fundamental differences in methodology may explain the discrepancies between our method’s results versus vendor-based GLS measurements,^17^ we speculate that DLS may enable better comparison with CMR feature-tracking measurements.

There are a few potential limitations to our methodology. Given the dependence on appropriate deep learning segmentation, we excluded studies for which deep learning failed. Not all cases were able to be accurately contoured, particularly if the video in question did not show all segments of the left ventricle or the lateral wall moved outside of the ultrasound sector during a cardiac cycle, which was the case in many segmentation failures. Inherent differences in methodology are present: our proof-of-concept study relies on the A4C view alone, whereas standard 2D GLS uses all three apical views, and 3D GLS uses 3D acquisitions. However, the measurements are comparable, and the methodology and the codebase are easily generalizable to other views.

Potential additional benefits of our pipeline are derived from the DLS measurement as a mathematical extension of a semantic segmentation model. In clinical practice, speckle-tracing algorithms often do not appropriately track the motion of the LV. The direct measurement and identification of the endocardial contour are both easily understandable and visually assessable for sources of error. Our algorithm enables rapid retrospective batch analysis of echocardiographic images, which may have applications in both research and clinical workflow while eliminating human-based measurement variance. Additionally, this pipeline could be adapted to other semantic segmentation models, allowing generalization to calculate right ventricular or atrial strain.

In conclusion, we present a deep-learning derived strain measurement based on deep learning derived endocardial contour. The results show that this measurement can be performed reliably with low variance and within the range of standard measurements. The DLS method is rapid, consistent, vendor-agnostic, publicly available, and robust across a wide range of image qualities.

## Supporting information

STARD

## Data Availability

Original training are available online at https://echonet.github.io/dynamic/
Other data is available upon reasonable request to authors

